# The Epidemiology of COVID-19 and applying Non Pharmaceutical interventions by using the Susceptible, Infectious Recovered epidemiological Model in Pakistan

**DOI:** 10.1101/2020.05.08.20095794

**Authors:** Abdul Wahid, Amjad Khan, Qaiser Iqbal, Asad Khan, Nazar Mohammad

## Abstract

**Introduction:** The COVID-19 is caused by the virus known as sever acute respiratory syndrome corona virus 2 (SARS-CoV-2) having the common symptoms such as Flue, fever, dry cough and shortness of breath. The first case was reported in WUHAN city china in December 2019 and it spread to the whole world, WHO declared as world pandemic on 11^th^ march 2020.

**SIR Epidemiological Model:** The first case in Pakistan was confirmed on 26^th^ Feb 2020 as by the 8^th^ April 2020 the total no of confirmed cases 4187 with 58 deaths and 467 recoveries throughout the country. The upcoming situation of the COVID-19 in Pakistan is forecasted by using SIR epidemiological, which is one of the mathematical derivative models with great accuracy rate prediction used for infectious disease. This model was introduced in the early 20^th^ century.

**Results:** Pakistan is will be having a heavy burden of patients 80000 plus infected patients 45000 recoveries 10000 hospitalized 3000 ICU and 800 plus deaths in the next 20 days. A complete lock down, social distancing and imposing curfew to keep every person at home can save Pakistan from a very huge number 1000000 infected patients with huge number of causalities with next 2 months.

## Introduction

The Coronaviruse disease 2019 (COVID-19) is caused by the virus known as sever acute respiratory syndrome corona virus 2 (SARS-CoV-2) having the common symptoms such as Flue, fever, dry cough and shortness of breath with complications to develop pneumonia and failure of the other vital organs which lead to death of the patient[1]. first case was identified in WUHAN city of Hubei province of China in December 2019[2].

The dramatic spread of virus through the whole world was declared COVID-19 pandemic by WHO on 11^th^ of march 2020[3]. More than 1363123 cases of COVID-19 have been reported in 209 countries and territories, resulting in approximately 76383 deaths. More than 293833 people have since recovered by 8^th^ April 2020[4].

The first case in Pakistan was confirmed on 26^th^ Feb 2020 as by the 8^th^ April 2020 the total no of confirmed cases 4187 with 58 deaths and 467 recoveries throughout the country[5]. Comparatively Pakistan with other countries such as UK, Iran, Italy, USA, Spain, Germany and other European countries, the no of positive patients of Pakistan are not as rapidly increasing as the no of patients of the other countries.

The early positive cases in Pakistan were received from the neighbor country Iran through the Toftan border thousands of persons (Zaireen) travel to Iran on monthly bases for their religious pilgrims (Ziarat) to the different cities of Iran[5]. The government of Pakistan didn’t allow the Zaireens to the local societies and kept them in Quarantines for 14 days with performing their COVID19 tests. All the schools Educational institutes were early closed for unknown period of time, the other markets, hotels and other crowded and rushed places were imposed a lock down for 20 days which may be extended for unknown period.

Pakistan is in the early stage of the COVID-19 pandemic. So it is very important to predict the COVID19 by using mathematical epidemiological models to predict the upcoming situation of the COVID-19 in Pakistan. The SIR epidemiological is one of the mathematical derivative models with great accuracy rate prediction used for infectious disease. This model was introduced in the early 20^th^ century by the scientist Kermack and McKendrick in 1927[6]. This model consist three compartments,

□ Susceptible Compartment (S)
□ Infected Compartment (I)
□ Recovered Compartment (R)

The whole population is placed in these three compartments by having same characteristics in same compartment. The patient infected will be removed from S compartment to I compartment and the patient who recover or die will be move from I compartment to R compartment.[7]

### 1. SIR Mathematical Model

This Mathematical Model has three compartments which are Susceptible (S), Infectious (I) and Recovered (R). The population of each compartment has its own characteristics the population of susceptible will remain susceptible unless comes in contact with infectious population and get infected will leave susceptible compartment and enter the infectious compartment and the infected population who recover or die will be removed from infectious compartment to recovered compartment.

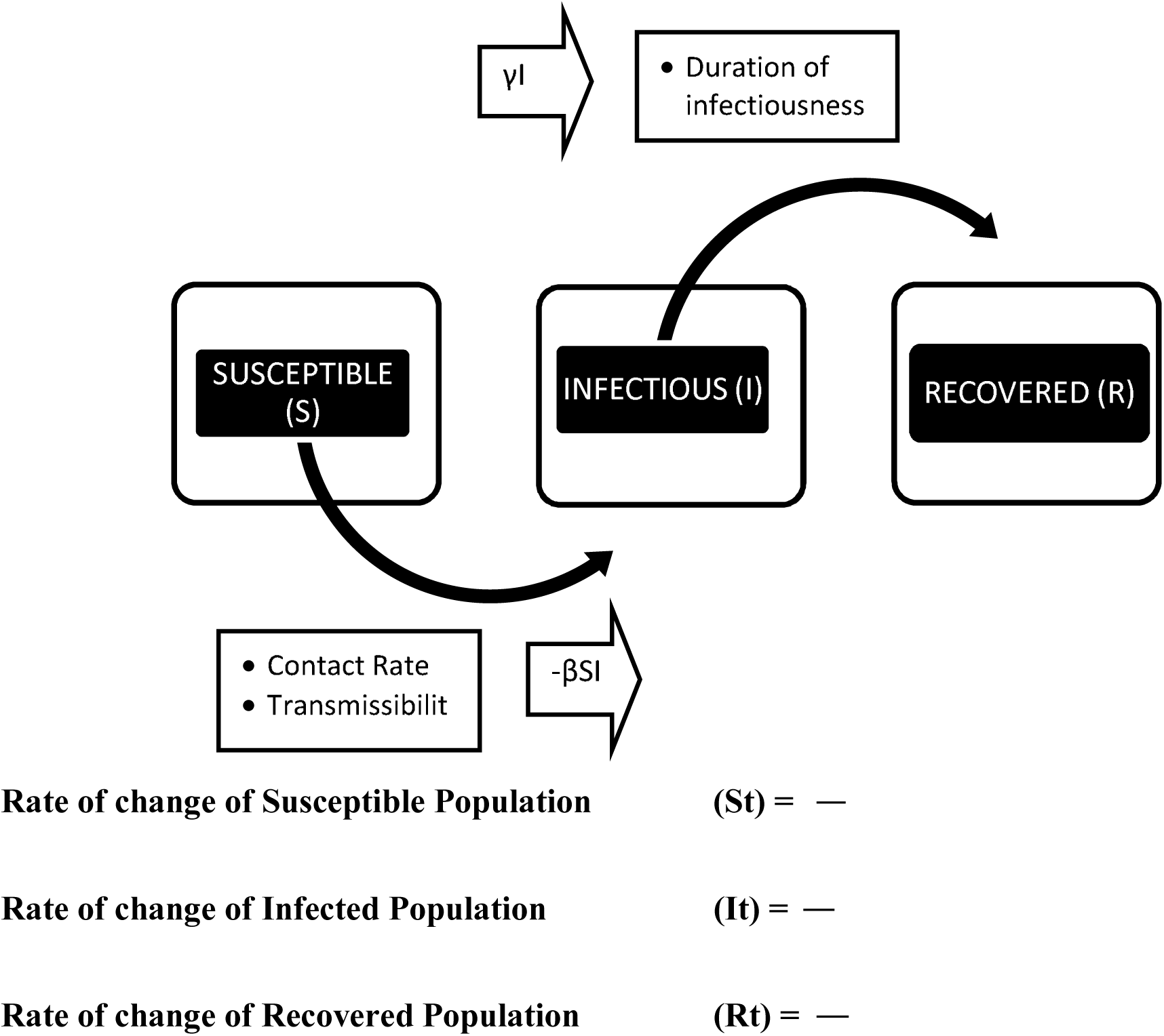

**Contact Rate:**

- The number of the susceptible people an infectious person contacts. Physical contact (Skin to skin contact) Non-physical contact (two-way conversation of 3+words in physical presence)
- Reflex mixing of the population.

**Transmissibility:**

- The probability that a contact between a susceptible person and infected person results in infection.

**Duration of Infectiousness:**

- The period of time that infected person can pass the infection.
- The inverse of the removal rate

**Reproduction number (Ro)**

- The number of secondary cases resulting from one case
- The product of Contact Rate x Transmissibility x Removal Rate

The COVID-19 has no approved pharmacological treatment and vaccine, so the non pharmacological intervention measures are adopted by all the countries worldwide such as social distancing, shut down of school, collages, universities and other education institutes, shut down of all the markets hotels and bazaars imposing a lock down a curfew with the slogan of STAY HOME SAVE LIVES[8]. Other measures of wearing mask using hand sanitizers increasing the hygiene can decrease the exposure to the virus.

So we take the contact rate of the population of susceptible compartment with infectious compartment as **β** for COVID-19.

- **β**: Determines how much the disease can spread due to exposure.
- The **β** value can be different for same virus but in different societies.
- A society with low contact rate will be having a lower **β** value and high contact rate will be having higher **β** value.

So the **β** value can be decreased by

- Border control by the Government
- Social Distancing decrease contact rate
- Stay at home
- Wearing Mask and Observe hygiene
- Developing the Vaccine Decreasing **β** value = Decreasing spread of infectious Rate So the contact rate is directly proportional to reproductive number of the COVID-19 increase in contact rate increases the reproductive number, decrease the contact rate decrease the reproductive no.

> ***Contact Rate ∝ Reproductive Number***
- **γ:** is a parameter expressing how much the infected population can be recovered in a specific period

Increase in **γ** value = Increase the Recovery Rate

The **γ** value can be increased by

- Better medicines

### 2. Results and Discussion

#### 2.1. Current situation of COVID-19 in Pakistan

Currently by 8^th^ April, 44896 tests (191/1M) are performed out of which 4322 COVID-19 were confirmed as positive and 63 deaths since 26^th^ Feb[9]. Comparing to other countries, USA 434927 positive cases and 14788 has deaths but the total test performed are 2209041 (6674/1M). Spain total no of tests 355000 (7593/1M) and confirmed cases are 148220 with 14792 deaths. In last 2 months above hundred thousand persons have been entered to Pakistan from different countries through airports from the European countries, from IRAN, AFGHANISTAN through the border gates out of these persons many were COVID-19 positive when their screening test were performed. (Figure 1A, Figure 1B) The demographic of COVID-19 positive patients in Pakistan are almost of all age categories both male and female as shown figure1 C.[5] (*Fig. 1A, 1B, 1C Here*) The whole population (220000000) of Pakistan is susceptible to COVID-19 infection, which was brought by very less number of people to the country. As this infection is going to spread the infectious period of the disease will start, so the number of infected will rise along with recovered cases. The rate at which susceptible population will become infected population depends on different factors: [10]

- At what speed the disease is going to spread naturally (unique to the virus)
- Contact rate of the population (unique to the society)
- How many tests performed for Detection? (unique to medical infra structure)

**Figure 1A.**
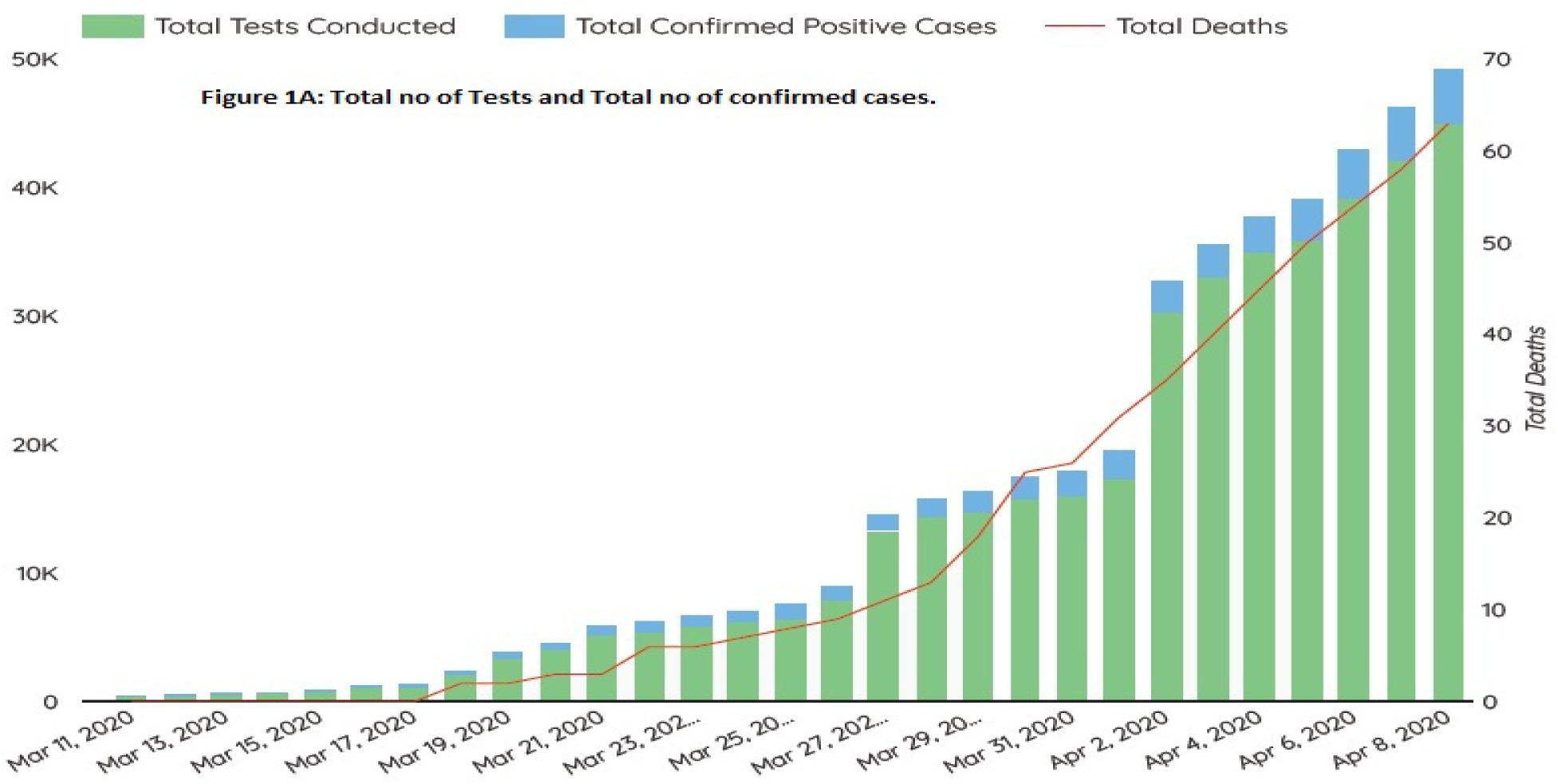

**Figure 1B.**
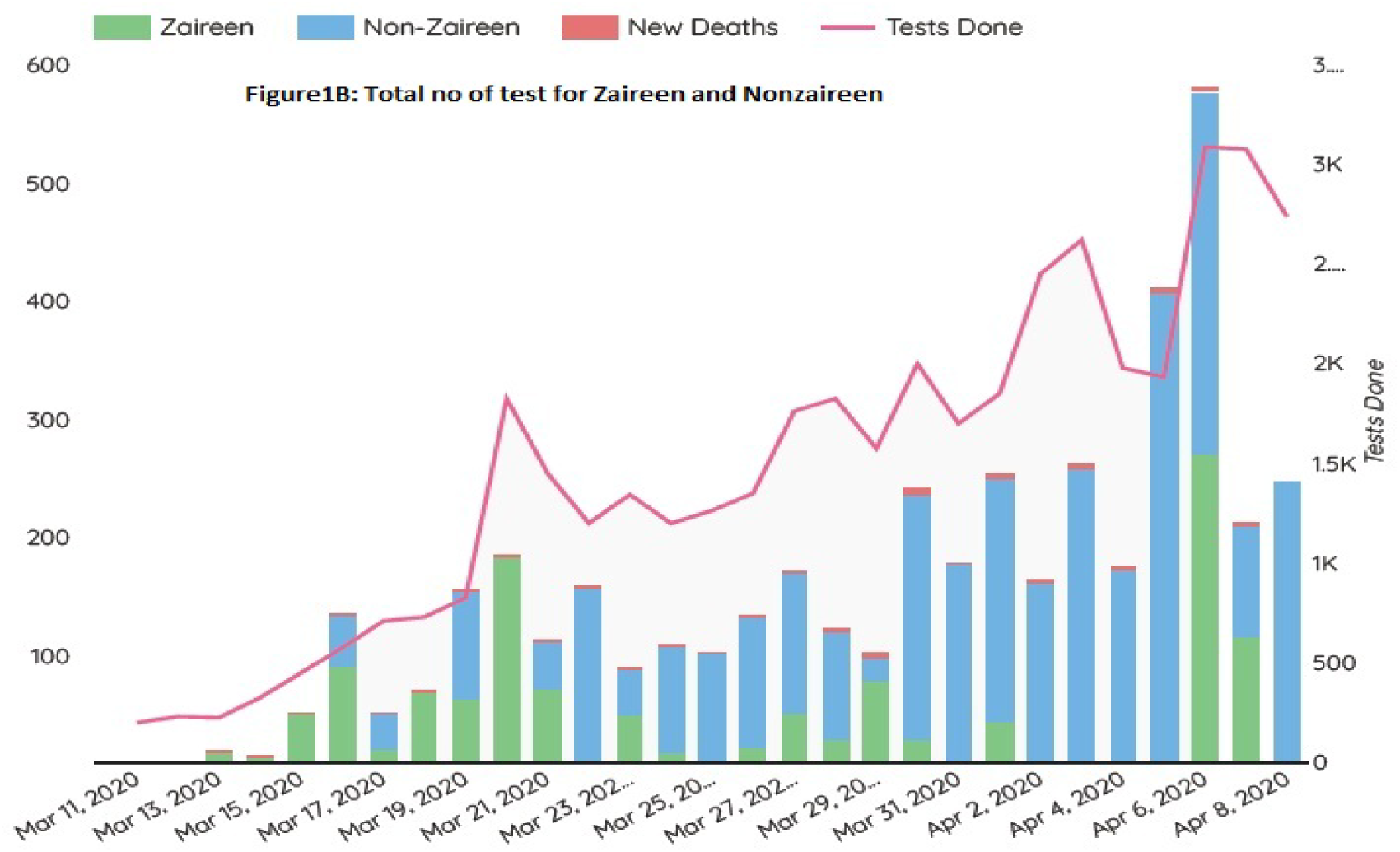

**Figure 1C.**
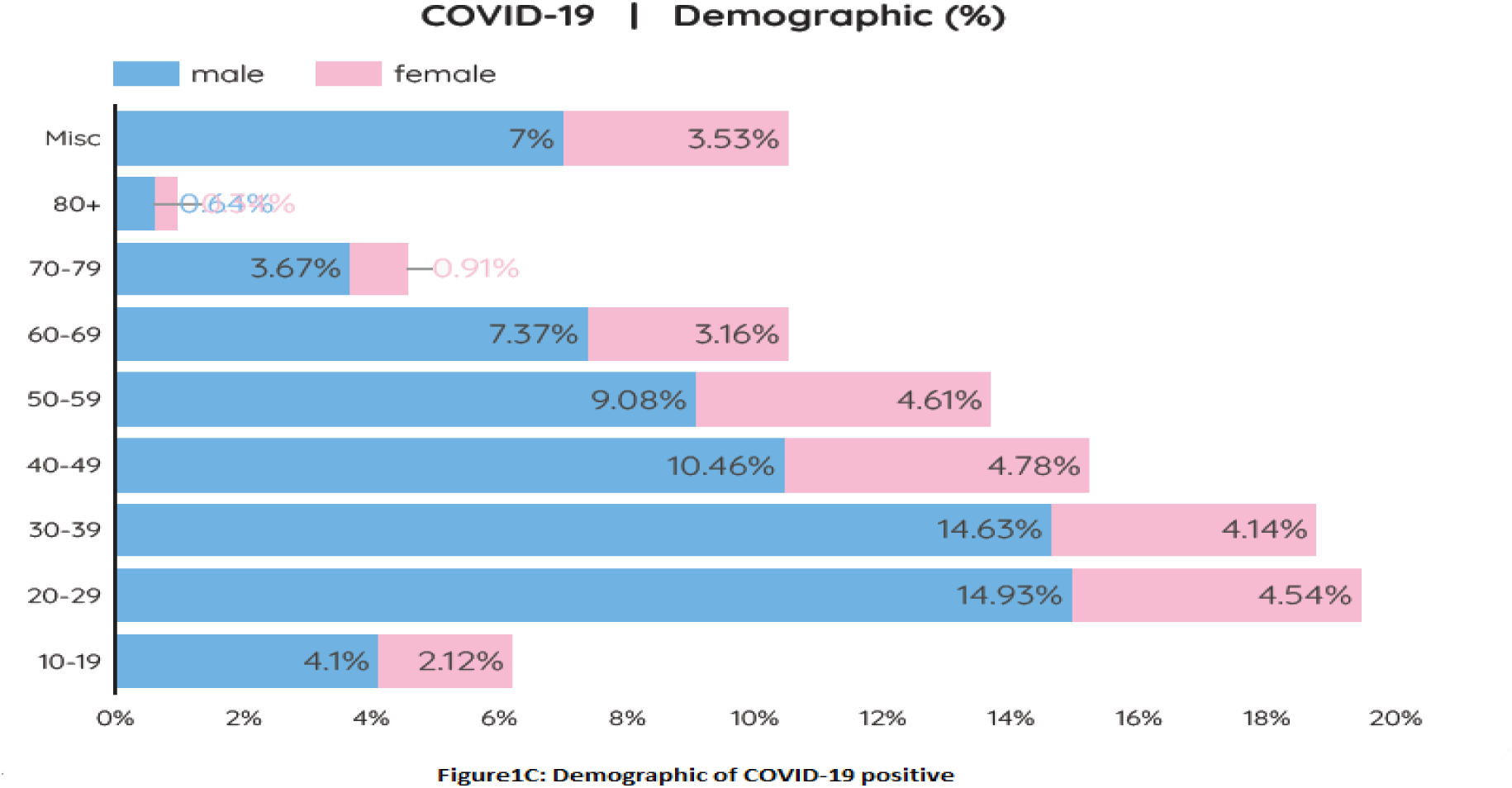

These are the factors which are responsible to the rate at which the susceptible population is going to be infectious population, and this parameter is known as **β**.

The Rate at which the people are going to recover is mostly related the pharmacological treatment and the unique to the virus, and this parameter is known as **γ** in the model.[11]

The Microsoft Excel sheet COVID-19 SIR Model created by Alex Hoyt, RN, PhD was used by changing the values according to this country population and contact rate.

By putting the values of Contact Rate, Transmissibility, Reproductive number, total susceptible, infected and recovered population in excel sheets the following graphs were obtained to predict the upcoming COVID-19 cases in Pakistan. The whole population of Pakistan (220000000) is susceptible to the COVID-19 and is expected to be infected but due to the very less no (2 cases on 26 –Feb) of the infected cases and due to the slow propagation of the infected cases the SIR model predicts the total susceptible, total Infected and total Recovered population for next 4 months i.e. 30/07/2020 as shown in figure 2A.

**Figure 2A.**
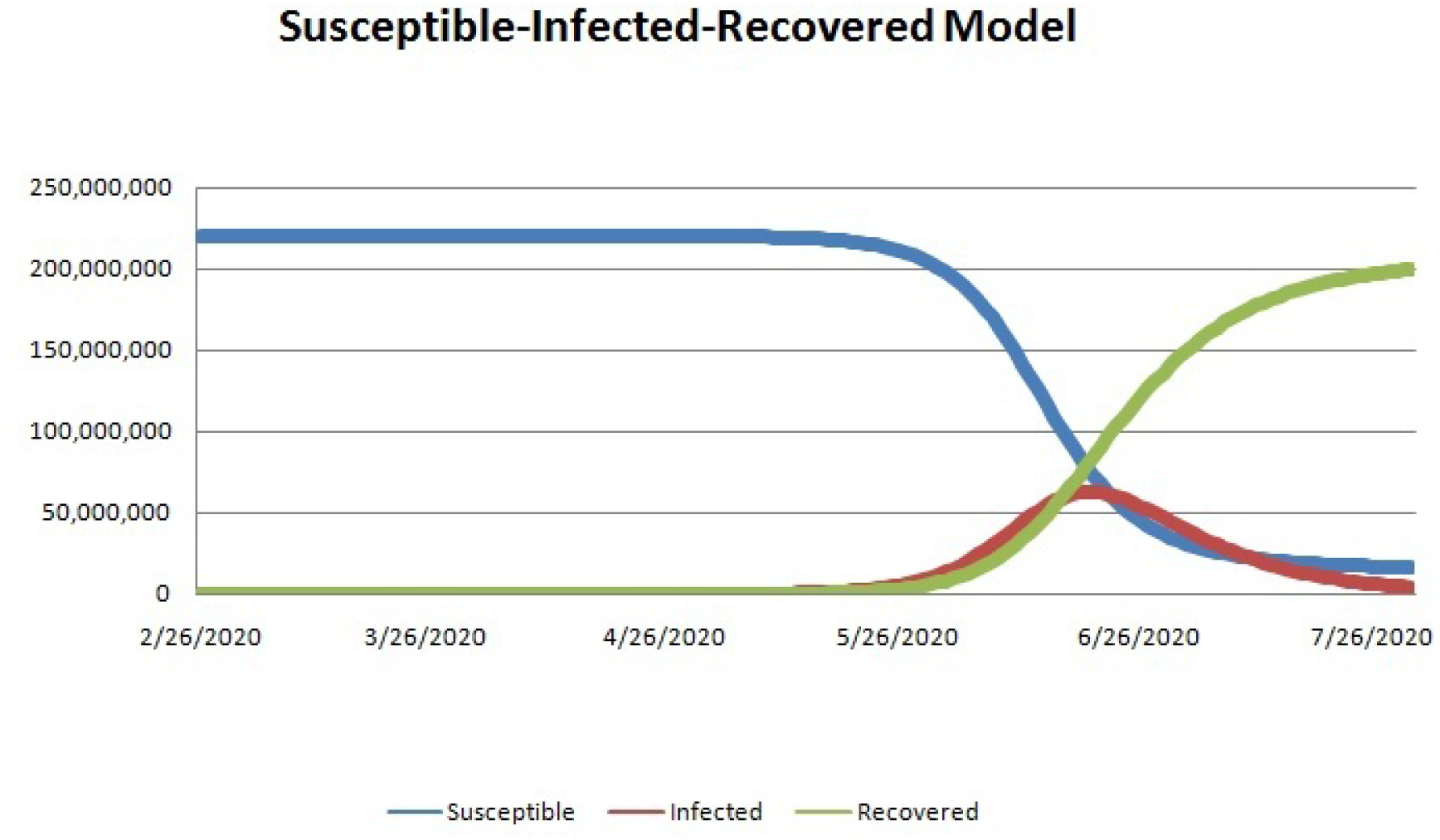

This model predicts that 50 million people will be infected in the upcoming four months if no proper measures are taken. These four months are going to be very challenging for Pakistan the state has to take every possible opportunity to combat the situation. The number of hospitalize patients are going to increase rapidly with increased fatality rate. *Figure 2A Here*

#### 2.2. The actual and forecast Cases

The actual cases which are confirmed by the government of Pakistan are from 26^th^ Feb 2020 as by the 8^th^ April 2020 the total infected cases 4187 with 58 deaths and 467 recoveries throughout the country[9]. The whole population is susceptible and according the present contact rate and transmissibility controlled by the government the predicted cases by the end of this month 30^th^ April 2020 the no of infected cases are going to reach 80000 plus with 800 plus deaths, 10000 plus hospitalized, 3000 plus ICU and 45000 plus recoveries as shown in the figure 2B. The no of the patient may increase or decrease which depends on contact rate and transmissibility. *Figure 2B Here*

**Figure 2B.**
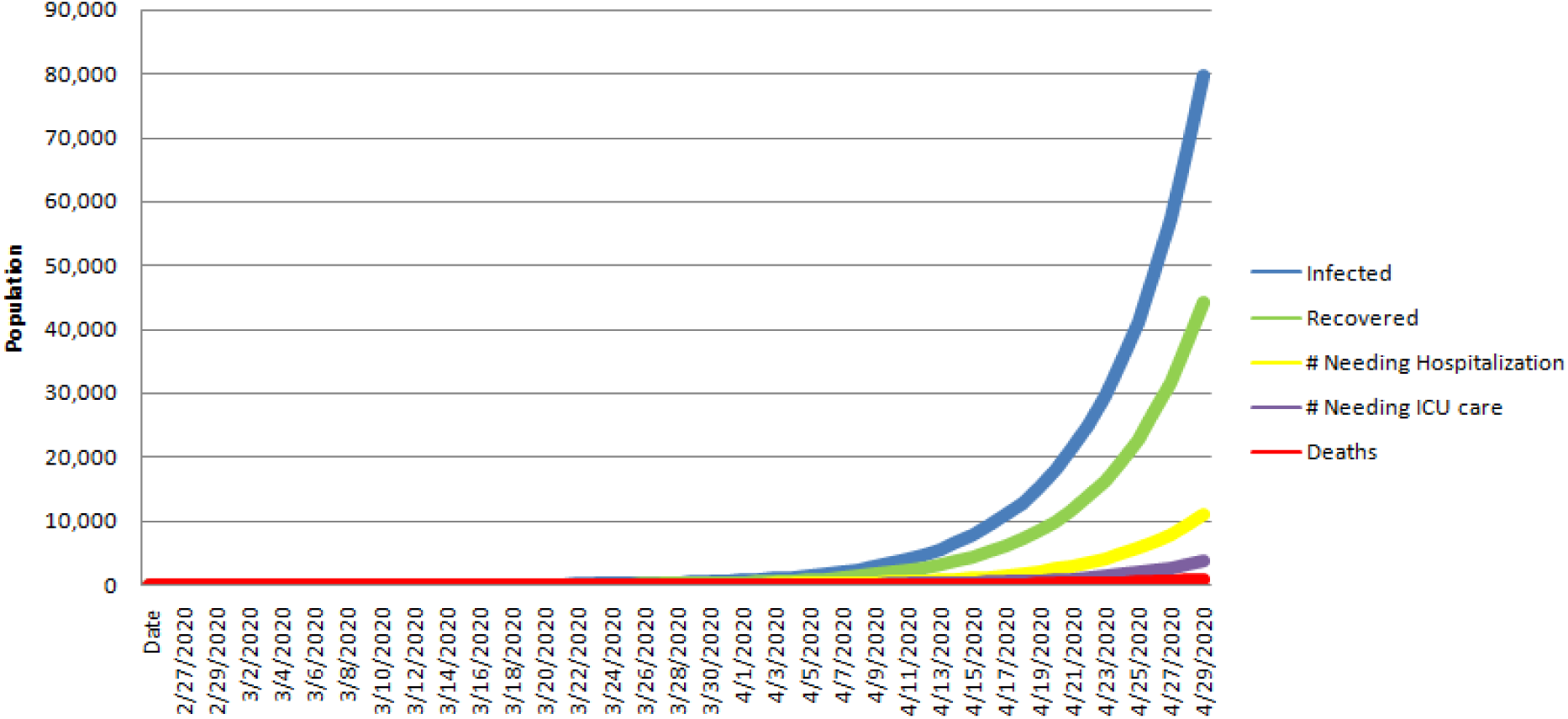

The health care system of Pakistan is not well equipped and developed to handle this large number of patients who will be hospitalized which will result in the collapse of the healthcare system in Pakistan[12] and the other emergencies and chronic disease may cause many deaths. To decrease these large numbers, the government should play its role through non-pharmacological interventions[13]. These interventions are decreasing the contact rate by isolation, quarantine, lock down or any other means that reduce the transmissibility rate which will reduce the number COVID-19 positive cases.[14]

#### 2.3. The Impact of Lock Down

The current situation of Pakistan is that all the educational institutes are closed till the 31^st^ May and all the markets, hotels, and other gatherings are prohibited till 20^th^ April. The Muslims offers 5 time prayer in 24 hours and majority of people offers their prayer in the Masque and a huge gathering on Friday (Juma Prayer) which are still not controlled by the government. The offices, work places, markets are closed but the public is not staying at home they are having huge gathering in play grounds, picnic points, sitting and wondering in bazaar and the huge gatherings for getting free food (Rashen) at many places in every city. The complete lock down and imposing curfew by restricting people to their houses can decrease the contact rate, transmissibility and ultimately can decreases the no of positive cases and death rate and increase recoveries.

When the exposure of the susceptible population is decrease to the infectious population so there will be decrease number positive cases vice versa as the exposure of susceptible and infectious population increase the number of positive patients will increase. So a complete lock down and imposing curfew, banning all religious and other gathering and restricting people to their homes can decrease the no of positive cases to limited no in two months. (*Figure 2C and Figure 2D Here*)

**Figure 2C.**
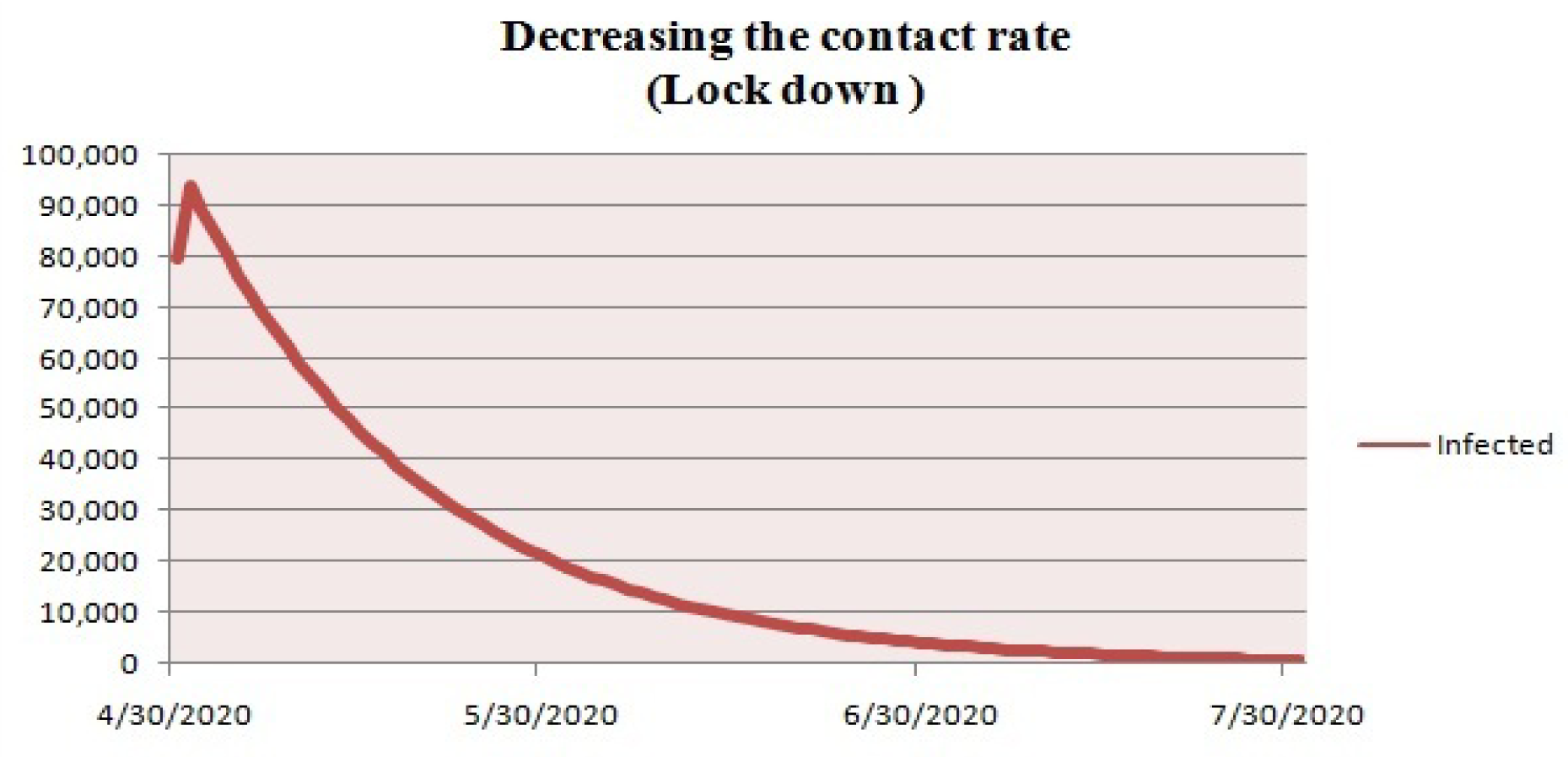

**Figure 2D.**
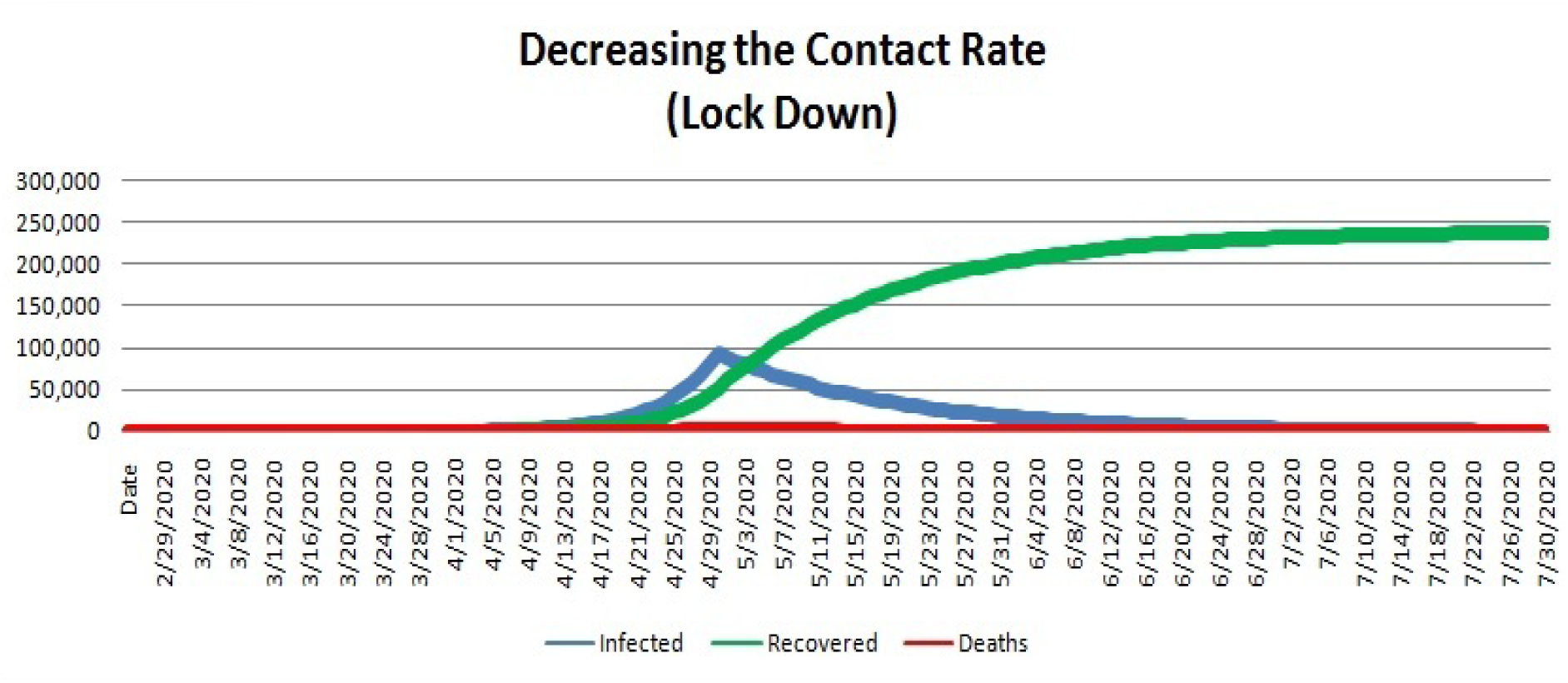

##### Limitation

- The SIR Model is closed system model the population of every compartment has its own characteristics.
- We have assumed the homogenous distribution of the Pakistan population that does not capture variations in population density or the urban-rural variations.
- We have assumed equal susceptibility to COVID-19, of all ages, gender and the having the co-morbidities
- The number of tests performed in Pakistan is very low as compare to the other countries so our absolute positive number might be low so we have assumed that increase the number of test will increase the number of absolute cases.

##### Advantages

- The SIR Model will Allow for easy modeling of epidemic
- This model can easily predict the no of infected, recovered, hospitalized, ICU and death for a period of time which can prepare the state for epidemics.
- This model can predict for both longer and shorter period of times
- Non Pharmacological interventions can easily be applied to the Model to see the impacts

### 3. Conclusion

Pakistan is currently in very early stage of the COVID-19. The growth rate of Pakistan is very low as compare to other countries. The SIR epidemic model shows that growth will increase exponentially and will reach 1000000 in the next 2 months and it will be difficult for Pakistan healthcare system to cope with the situation. This model also suggests that the Non-pharmacological Interventions (NPI) such as social distancing, complete lock down or imposing curfew, observing hygiene by washing hands, using hand sanitizers and wearing mask can slow down the propagation of the COVID-19 and can reduce the exposure of susceptible population to the infectious population.

Hence decreasing the contact rate and transmissibility can surely decline the graph for infections, hospitalizations, ICU admissions and ultimately the decrease in mortality rate up to 90% in the next two months.

## Data Availability

All data in this perspective will be made available.

---

**Table 1.** Values used for analysis.

|  |  |  |
| --- | --- | --- |
| **Contact Rate** | 14 | The contact rate has been found by taking the data from people of all ages and gender both rural and urban population through a questioner and calculated the mean. So NPIs we just decrease the contact rate according to the situations. |
| **Transmissibility** | 2.0% | The value used, 2.0%, was estimated by comparing the Reproduction Number ($R_0$) in community settings (1.4 to 3.9) with a normal contact rate whereas transmissibility and removal rate (inverse of duration of infectiousness) were assumed constant. |
| **Duration of Infectiousness** | 10 | Contact tracing data from 10 early cases in China showed the mean serial interval (time between successive cases) was 7.5 days with a SD of 3.4 days. A more recent estimate among 468 cases was 3.96 days with a 4.75 day SD. |
| **% Needing Hospitalization** | 13.8% | Among 44,672 confirmed COVID-19 cases in China, 13.8% required hospitalization. |
| **% Needing ICU Care** | 4.7% | Among 44,672 confirmed COVID-19 cases in China, 4.7% required ICU care. |
| **Mortality rate** | 1.5% | As of 8/04/2020, the average daily mortality rate of COVID19 is 1.5% |
| **Reproduction Number ($R_0$)** | | COVID-19 has been between 1.4 to 3.9. $R_0$ is also an indication of the effectiveness of community interventions. An $R_0$ less than 1 indicate transmission has stopped. |

## References

1. Xu, Z., et al., Pathological findings of COVID-19 associated with acute respiratory distress syndrome. The Lancet respiratory medicine, 2020. 8(4): p. 420–422.

2. Guan, W., et al., China Medical Treatment Expert Group for Covid-19. Clinical characteristics of coronavirus disease, 2019.

3. Cucinotta, D. and M. Vanelli, WHO Declares COVID-19 a Pandemic. Acta bio-medica: Atenei Parmensis, 2020. 91(1): p. 157.

4. worldometer, (https://www.worldometers.info/coronavirus/).

5. Pakistan, https://www.covid.gov.pk/covid19. 2020.

6. Kermack, W.O. and A.G. McKendrick, A contribution to the mathematical theory of epidemics. Proceedings of the royal society of london. Series A, Containing papers of a mathematical and physical character, 1927. 115(772): p. 700–721.

7. Chatterjee, K., et al., Healthcare impact of COVID-19 epidemic in India: A stochastic mathematical model. Medical Journal Armed Forces India, 2020.

8. Naresh Dhanwant, J. and V. Ramanathan, Forecasting COVID 19 growth in India using Susceptible-Infected-Recovered (SIR) model. arXiv, 2020: p. arXiv: 2004.00696.

9. Pakistan, https://www.nih.org.pk/novel-coranavirus-2019-ncov/. 2020.

10. Peng, L., et al., Epidemic analysis of COVID-19 in China by dynamical modeling. arXiv preprint arXiv:2002.06563, 2020.

11. Simha, A., R.V. Prasad, and S. Narayana, A simple Stochastic SIR model for COVID 19 Infection Dynamics for Karnataka: Learning from Europe. arXiv preprint arXiv:2003.11920, 2020.

12. Khalid, F. and A.N. Abbasi, Challenges faced by Pakistani healthcare system: Clinician’s perspective. 2018.

13. Lai, S., et al., Effect of non-pharmaceutical interventions for containing the COVID-19 outbreak: an observational and modelling study. medRxiv, 2020.

14. Krr, G. and F. Casella, Non-Pharmaceutical Interventions (NPIs) to Reduce COVID-19 Mortality. Available at SSRN 3560688, 2020.

